# Community-onset urinary tract infection in females in the context of COVID-19: a longitudinal population cohort study exploring case presentation, management, and outcomes

**DOI:** 10.1101/2023.07.16.23292705

**Authors:** Nina J Zhu, Benedict Hayhoe, Raheelah Ahmad, James R Price, Donna Lecky, Monsey McLeod, Elena Farren, Timothy M Rawson, Emma Carter, Alison H Holmes, Paul Aylin

## Abstract

**Background:** COVID-19 affected the epidemiology of other infectious diseases and how they were managed. Urinary tract infection (UTI) is one of the most common infections treated in the community in England. We investigated the impact of the COVID-19 pandemic on UTI primary care consultations and outcomes in female patients.

**Methods and findings:** We analysed General Practice (GP) consultation and hospital admission records using the Whole Systems Integrated Care (WSIC) data in North West London between 2016 and 2021. We quantified the changes in UTI GP consultation rates using time series analysis before and during the pandemic. We assessed the outcomes of UTI, measured by subsequent bacteraemia and sepsis within 60 days, for consultations delivered face-to-face or remotely, with or without diagnostic tests recommended by the national guidelines, and with or without antibiotic treatment. Between January 2016 and December 2021, we identified 375,859 UTI episodes in 233,450 female patients. Before the COVID-19 pandemic (January 2016 – February 2020), the UTI GP consultation rate stayed level at 522.8 cases per 100,000 population per month, with a seasonal pattern of peaking in October. Since COVID-19, (March 2020 – December 2021), monthly UTI GP consultations declined when COVID-19 cases surged and rose when COVID-19 case fell. During the pandemic, the UTI consultations delivered face-to-face reduced from 72.0% to 29.4%, the UTI consultations with appropriate diagnostic tests, including urine culture and urinalysis, reduced from 17.3% to 10.4%, and the UTI cases treated with antibiotics reduced from 52.0% to 47.8%. The likelihood of antibiotics being prescribed was not affected by whether the consultation was delivered face-to-face or remotely but associated with whether there was a diagnostic test. Regardless of whether the UTI consultation occurred before or during the pandemic, the absence of antibiotic treatment for UTI is associated with a 10-fold increase in the risk of having bacteraemia or sepsis within 60 days, though the patients who consulted GPs for UTI during the pandemic were older and more co-morbid. Across the study period (January 2016 – December 2021), nitrofurantoin remained the first-line antibiotic option for UTI. The percentage of non-prophylactic acute UTI antibiotic prescriptions with durations that exceeded the guideline recommendations was 58.7% before the pandemic, and 49.4% since. This led to 830,522 total excess days of treatment, account for 63.3% of all non-prophylactic acute antibiotics prescribed for UTI. Before the pandemic, excess antibiotic days of UTI drugs had been reducing consistently. However, this decline slowed down during the pandemic. Having a diagnostic test was associated with 0.6 less excess days of antibiotic treatment.

**Conclusions:** This analysis provides a comprehensive examination of management and outcomes of community-onset UTI in female patients, considering the changes in GP consultations during the COVID-19 pandemic. Our findings highlighted the importance of appropriate urine testing to support UTI diagnosis in symptomatic patients and initiation of antibiotic treatment with appropriate course duration. Continued monitoring is required to assess the overall impact on patients and health systems from the changed landscape of primary care delivery.

## Introduction

Urinary tract infections (UTI), defined by a combination of clinical features and the presence of bacteria in the urine, accounted for 1-3% of all general practice (GP) consultations in England [1]. Three-quarters of women will have a UTI in their lifetime, with *Escherichia coli* (*E. coli*) being the most common causative pathogens, identified in more than 70% of the cases [2]. The clinical spectrum of UTI ranges from mild urinary symptoms through to urosepsis, and in some cases, can lead to bacteraemia that requires hospitalisation. UTI is the second most common reason for antibiotics to be prescribed in primary care in England [3]. However, up to 50% of antibiotic prescriptions for UTI are inadequate [4]. Inappropriate management of UTI includes both under- and over-treating with antibiotics. Under-treating UTI, meaning absence or delay of appropriate antibiotic treatment, significantly increases the risk of developing BSI, particularly in elderly patients with lower UTI [3,5]. In contrast, over-treating UTI, including using antibiotics when not required or using antibiotics with a prolonged duration, accelerates the emergence and transmission of antimicrobial resistance (AMR) in the long-term [6]. Up to 70% of females with urinary tract symptoms were found not to have a UTI confirmed microbiologically using routine urine culture [8]. Urine culture and urinalysis are critical to support prudent prescribing practices [7]. However, the COVID-19 pandemic has brought great challenges to infection diagnosis and management in primary care [9]. During UK’s first surge of COVID-19 cases, non-emergency services were suspended, and most GP consultation switched from face-to-face appointments to remote delivery through video or telephone call. Between April to May 2020, the weekly rate of UTI GP consultations reduced substantially from a 5-year average of 30 to 35 to less than 10 per 100,000 population, with fewer urine dipstick tests performed and fewer urine samples requested for culture [10]. The COVID-19 pandemic also has had impact on primary care antibiotic consumption. A significant reduction in GP antibiotic prescribing was observed across the first two COVID-19 waves in North West London, including 16.9% decrease in UTI antibiotics (nitrofurantoin, trimethoprim) [11], giving rise to the possibility that there were missed case presentation of patients which were untreated. Missing UTI treatment is especially risky in the elderly population who have a greater likelihood of developing complications, such as BSI or sepsis [10,12]. There is an urgent need to understand the scale of the impact of COVID-19 on how UTI cases were managed in the community, and the subsequent patient outcomes. In this analysis, we aimed to analyse the trends in primary care UTI diagnosis in female patients in North West London, and assessed the potential influence on antibiotic treatment decisions and outcomes from changed patient mix and GP consultation delivery methods since COVID-19.

## Methods

### Ethical approval

This study was approved by the Imperial Academic Health Science Centre (AHSC) COVID Research Committee, the COVID-19 NWL Data Prioritisation Group, and the Discover Research Advisory Group (DRAG), which jointly provides a governance mechanism.

### Data and study population

We used de-identified individual-level linked datasets from Imperial NIHR Biomedical Research Centre (BRC) Clinical Analytics, Research and Evaluation (iCARE) high performance analytics environment [13], which hosts primary care records from Whole Systems Integrated Care (WSIC) and secondary care records from NHS Secondary Uses Service (SUS) [14], covering more than 2 million patients in NWL [15]. In this analysis, we included female patients who consulted a general practitioner for a new episode of community-onset lower UTI between 01 January 2016 and 31 December 2021, as identified by all primary care diagnosis codes for suspected or uncomplicated UTI (Supplementary Material Table S1). If a patient had multiple UTI consultations, all consultations within a static 60-day window were grouped into one UTI episode (*e.g.,* if a patient had UTI GP consultations on 01 June 2018, 05 June 2018, and 15 August 2018, this patient was considered to have 2 UTI episodes onset on 01 June 2018 and 15 August 2018 respectively). We extracted GP antibiotic prescriptions by identifying Antibacterial drugs listed in British National Formulary (BNF) [16]. We also extracted all the bacteraemia-related GP consultations or hospital admissions, identified by all primary care or hospital diagnosis codes for bacteraemia or sepsis (Supplementary Material Table S2, S3).

Patients were excluded from analysis if: gender-identify which was not defined as female, *or* had a GP UTI diagnosis before 01 January 2016 (i.e., only include UTI episodes which were newly onset during the study period), *or* had UTI that originated in hospital (*i.e.,* removed any patients identified with ICD-10 coded hospital diagnosis before or at the same time of the initial UTI GP diagnosis).

### Descriptive analysis

We analysed UTI outcomes measured by 60-day bacteraemia, and sepsis, and all-cause mortality, stratified by before or during the COVID-19 pandemic (pre-pandemic period: from January 2016 to February 2020; pandemic period: from March 2020 to December 2021), patient characteristics, and how UTIs were diagnosed and managed. Patient risk factors include age at episode start (under 16 years old; 16 – 64 years old; above 64 years old), quintile of socioeconomic status (Index of Multiple Deprivation [IMD] 2019, 1 = most deprived, 5 = least deprived), ethnicity (non-white; white; unknown); number of long-term conditions (with 0 conditions; with 1 condition; with 2 or more conditions); the Cambridge Multimorbidity Score (CMS) (weighted score combining 20 conditions based on prevalence and effect size for mortality or particular outcome prediction) (Supplementary Material Table S4); recent hospitalisation (being discharged 30 days before UTI onset); COVID-19 status (whether the UTI episode was associated with a positive SARS-CoV-2 test 14 days before or after the UTI onset); and residential status (whether a resident in long-term care facilities). To account for the excess mortality potentially caused by COVID-19 during the pandemic, we re-calculated the 60-day all-cause case fatality rate after excluding those who had laboratory-confirmed SARS-CoV-2 infection. To address the high collinearity between living in a care home and age, we used the matched case-control approach by comparing the treatment for each care home resident against a randomly matched patient who had the same age and did not live in a care home. We also assessed the factors that might potentially influence antibiotic prescribing and subsequent patient outcomes, including consultation methods (remote consultation via telephone, video, letter, email, or text message; face-to-face consultation in GP surgeries, out-of-hours clinics, walk-in centres, or via home visit), whether there was a diagnostic test in line with the National Institute for Health and Care Excellence (NICE) guideline (NG109) for UTI management (for patient aged under 16: urine dipstick, urine culture, or urinalysis; for patient aged 16 and above: urine culture or urinalysis) [2]. We considered a UTI consultation supported by testing if there was either the order of a test or the results of a test documented within 72 hours following the initial GP consultation to allow for laboratory turnaround time of urine microbiology [17]. We also categorised the UTI episodes by treatment (had antibiotics: if there was a prescription from the same day as the initial UTI GP consultation up to 7 days following the initial UTI GP consultation considering back-up prescribing; had not been treated with antibiotics). We presented the durations of antibiotic prescriptions and the number of excess antibiotic days, defined as the total number of days beyond the recommended duration in the guidelines [2]. We excluded prescriptions that were explicitly coded as a repeat prescription, for recurrent cases, and for pregnant women (for whom prophylactic prescribing for recurrent UTI and asymptomatic bacteriuria in pregnancy was common) [18].

### Statistical analysis

This study includes two statistical analyses. First, a multi-phase interrupted time-series analysis to compare trends of GP UTI consultations rates among female patients in North West London before and since COVID-19. We constructed monthly time series for UTI episode counts per 100,000 female population, using Office for National Statistics’ (ONS) Mid-Year Population Estimates as a denominator. We defined the following six stages: pre-pandemic (Jan 2016 - March 2020); COVID-19 wave 1 (March 2020 - May 2020); between wave 1 and 2 (June 2020 – October 2020); COVID-19 wave 2 (November 2020 – February 2021); between wave 2 and 3 (February 2021 – June 2021); and wave 3 (July 2021 – Feb 2022) (Figure 1) [19].

**Figure 1.**
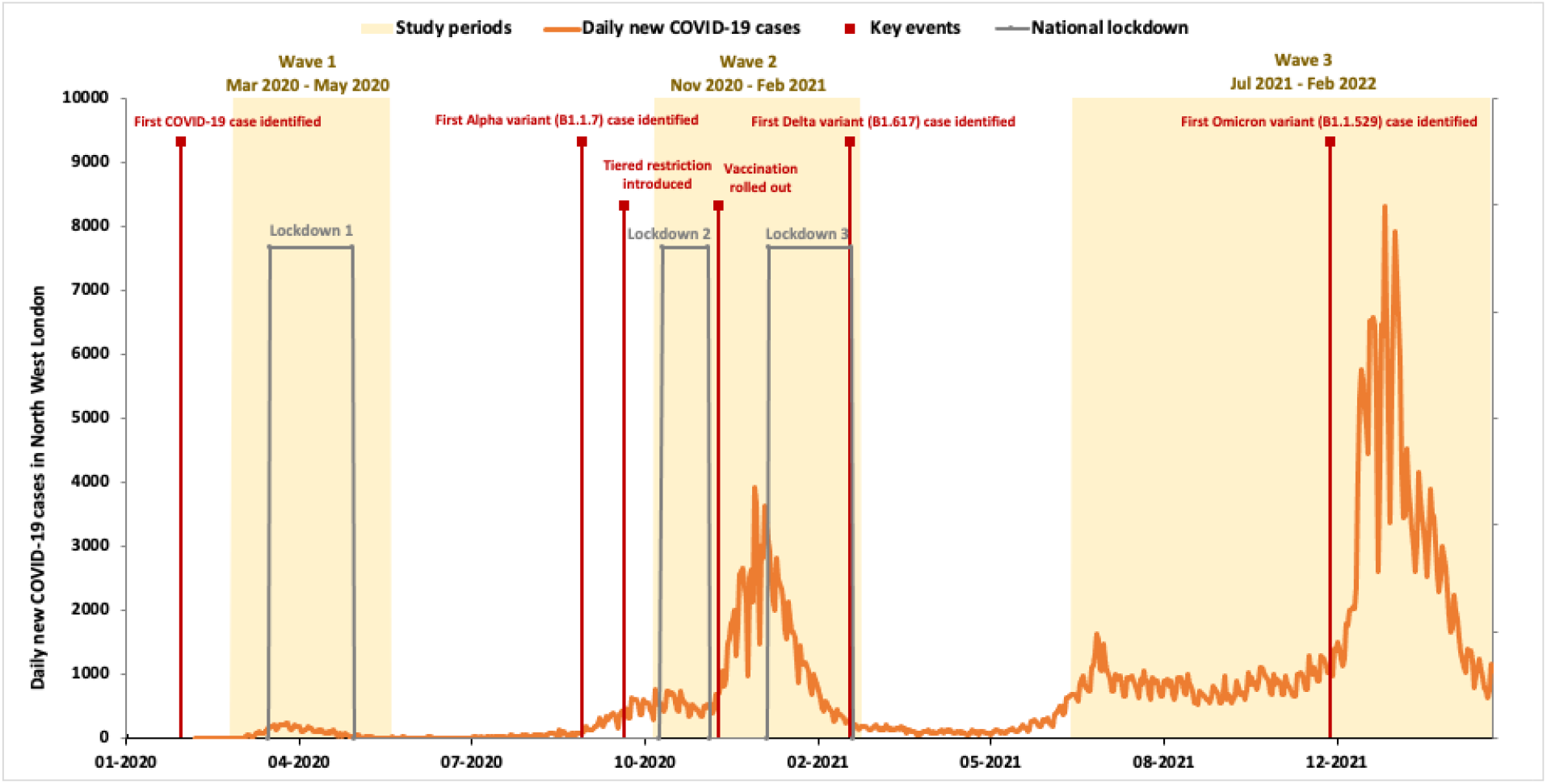
Defined COVID-19 pandemic waves for the interrupted time-series analysis

Our analysis relied on ordinary least squares (OLS) regression based on autoregressive integrated moving-average methods [20]. The regression assumes the following form (Linden and Adams 2011):. We performed Cumby-Huizinga test to specify the lag order when autocorrelation is present for the residual term. Secondly, we constructed the monthly time series of bacteraemia and sepsis diagnosed by GP or in hospitals (including those without prior GP diagnosis of UTI) for the time period between January 2016 to February 2021. This allowed us to assess whether there has been an increase in GP consultation or hospitalisation due to bacteraemia and sepsis in female patients, which could be explained by those UTI cases which were undiagnosed and untreated during COVID-19 surges. Second, we undertook a univariable analysis comparing patients with and without immediate antibiotic treatment for each included variable. To assess the difference between variables, we used the χ2 test for categorical variables, the Kruskal-Wallis H test for continuous variables. A p-value below 0.05 was considered statistically significant. We presented regression coefficient, unadjusted odds ratio (OR), and adjusted odds ratio (aOR) with 95% confidence intervals (95% CI).

## Results

From the WSIC database we extracted 1,183,064 GP coded UTI consultations (grouped into 546,969 new-onset UTI episodes) between 01 January 2016 to 31 December 2021. After applying our exclusion criteria and removing all duplicates, our analytical sample included 375,859 distinct UTI episodes.

### Epidemiological patterns of urinary tract infections (UTIs): before and during the COVID-19 pandemic

The monthly UTI GP consultation rate before COVID-19, between January 2016 and February 2020, was stable at 523 cases per 100,000 population per month. When monthly variation was considered, seasonality was observed with peak UTI GP consultation rate in October every year (p = 0.118). Given the pre-pandemic trajectories, we estimated the monthly UTI GP consultation rate under the hypothetical scenario of absence of the COVID-19 pandemic (Figure 2). During COVID-19 wave 1 (March 2020 - May 2020), UTI GP consultation rate deviated from the historical pattern and changed to a declining trend, decreasing by 24.2 episodes per 100,000 population per month (p = 0.000). Between wave 1 and 2 (June 2020 – October 2020), the UTI GP consultation rate experienced a sharp upward trend, increasing by 17.6 episodes per 100,000 population per month (p = 0.000). The pattern of UTI GP consultation rate decreased when COVID-19 cases increased, persisted during the later stages of the pandemic though the changes during COVID-19 wave 2 (November 2020 – February 2021) and between wave 2 and 3 (February 2021 – June 2021) (Figure 2, Supplementary Material Table S5).

**Figure 2.**
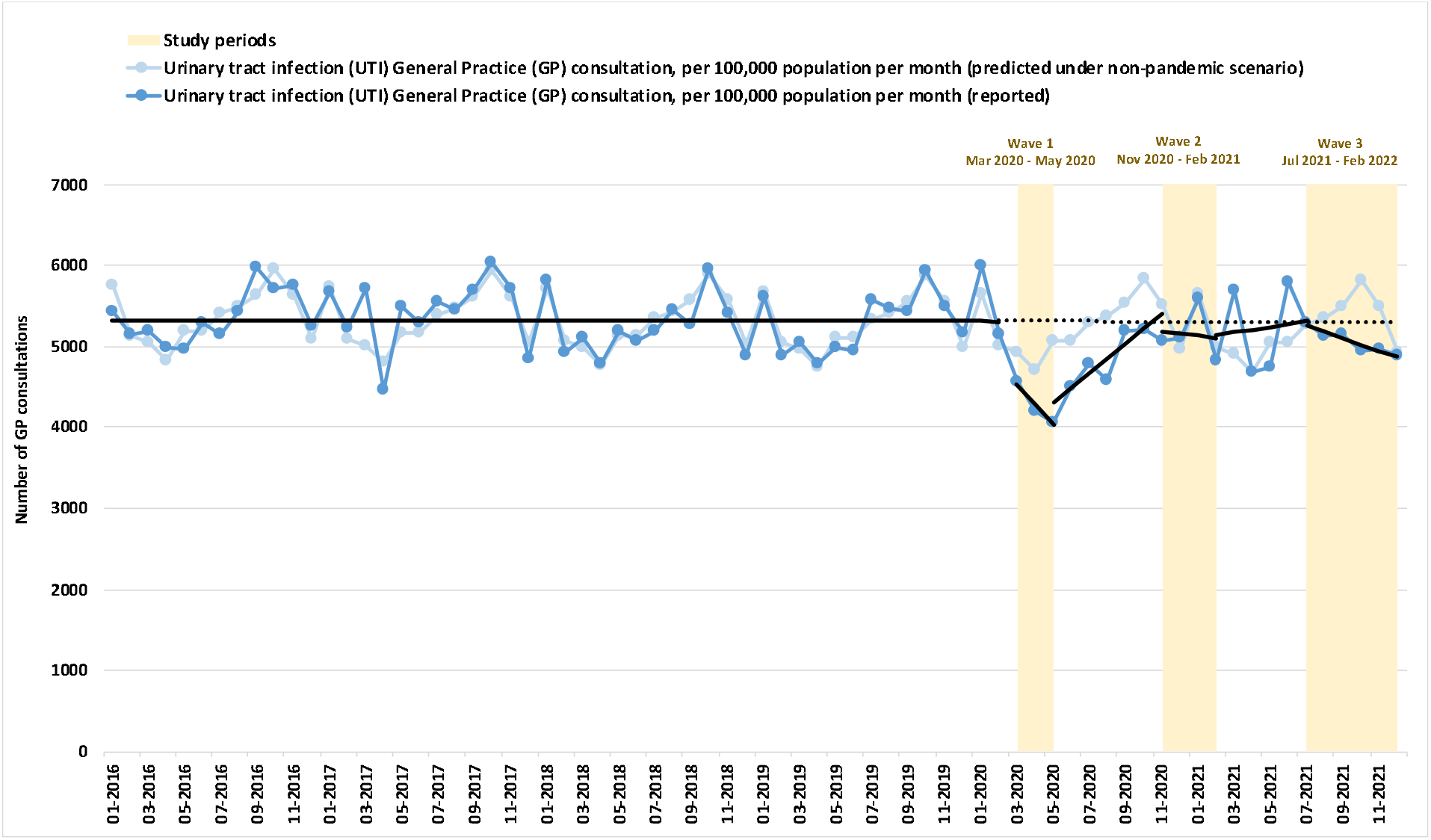
Number of urinary tract infection (UTI) General Practice (GP) consultations per 100,000 population per month

### Patient characteristics of urinary tract infection (UTI): before and during the COVID-19 pandemic

Our study included 233,450 unique female patients, their mean age at the time of the initial UTI diagnosis (or at the event of the first episode if had recurrent UTIs) of the study cohort was 41.6 years (SD = ±22.2 years). 22.0% (n = 51,256) of the patients had multi-morbidities (co-occurrence of two or more chronic diseases), 25.9% (n = 43,956) of the patients had recurrent UTIs. 0.6% (n = 1,422) patients were pregnant when UTI was diagnosed. 1.5% (n = 3,594) patients once lived in care homes, they had a mean age of 82.7 years (SD = ±12.8 years). We identified 375,859 distinct UTI episodes among the study cohort. We present the patient characteristics based on when the UTI episode was identified (Table 1). There were 266,946 UTI episode occurred before COVID-19, the patients had the mean age of 44.8 years, 30.9% (n = 82,437) UTI were from patients with multi-morbidities. Since March 2020, 108,913 UTI episodes were identified from patients with an older mean age (45.3 years), and higher co-morbidities (33.7% (n = 36,748) UTI from patients with multi-morbidities). 84,560 patients consulted GP for UTI during the pandemic period, of which 13.7% (n = 11,600) had laboratory confirmed SARS-CoV-2 infection.

**Table 1.**
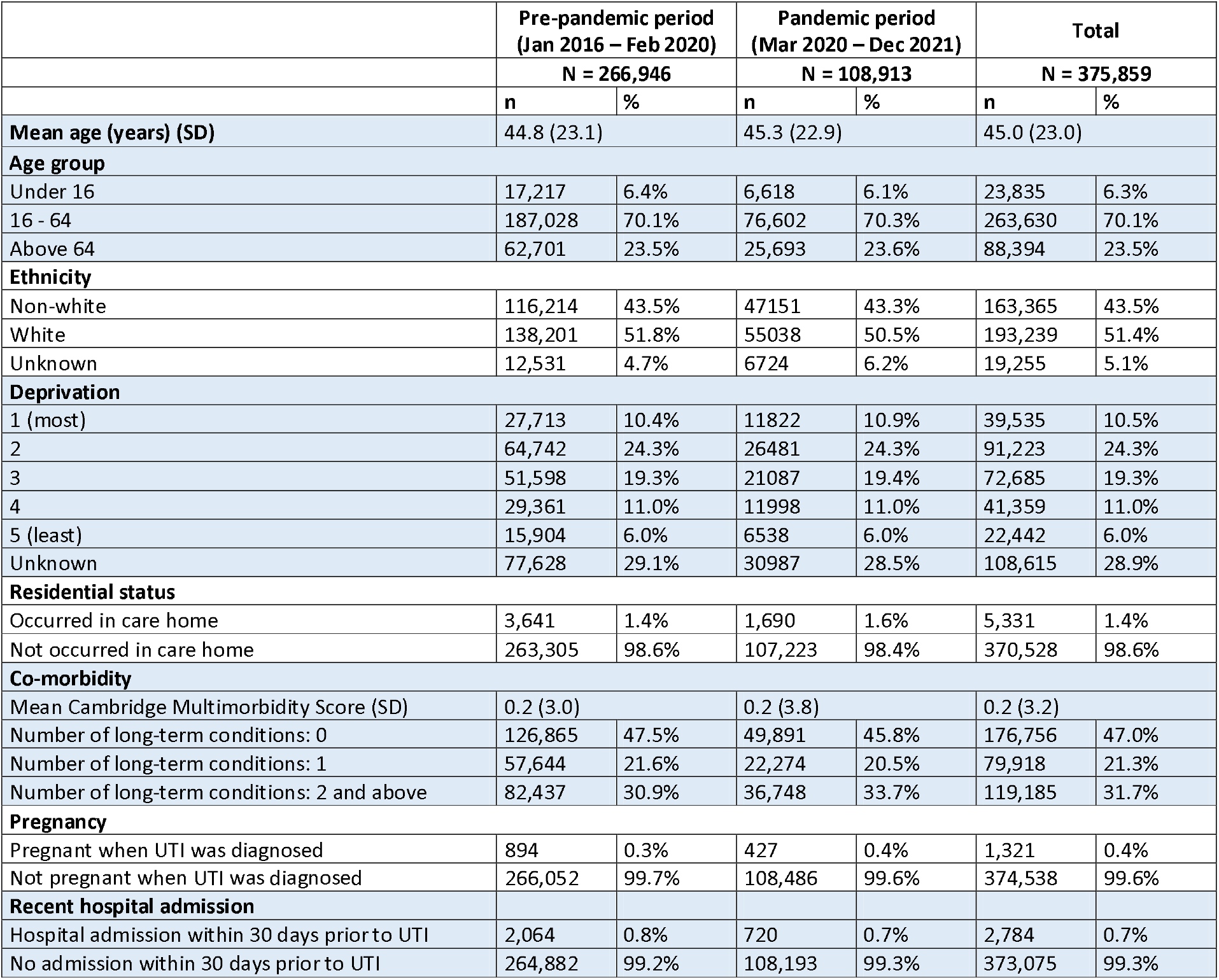
Patient characteristics of urinary tract infection (UTI) General Practice (GP) consultations

During COVID-19 surges, UTI cases decreased with the most significant decline in the patients aged 18 – 64 (Figure 3(a)). Overall, the patients who consulted a GP for UTI were older and had worse health, measured by both multi-morbidity score and number of chronic conditions, during the pandemic waves (Kruskal-Wallis H test, p = 0.001) (Figure 3(a)).

**Figure 3.**
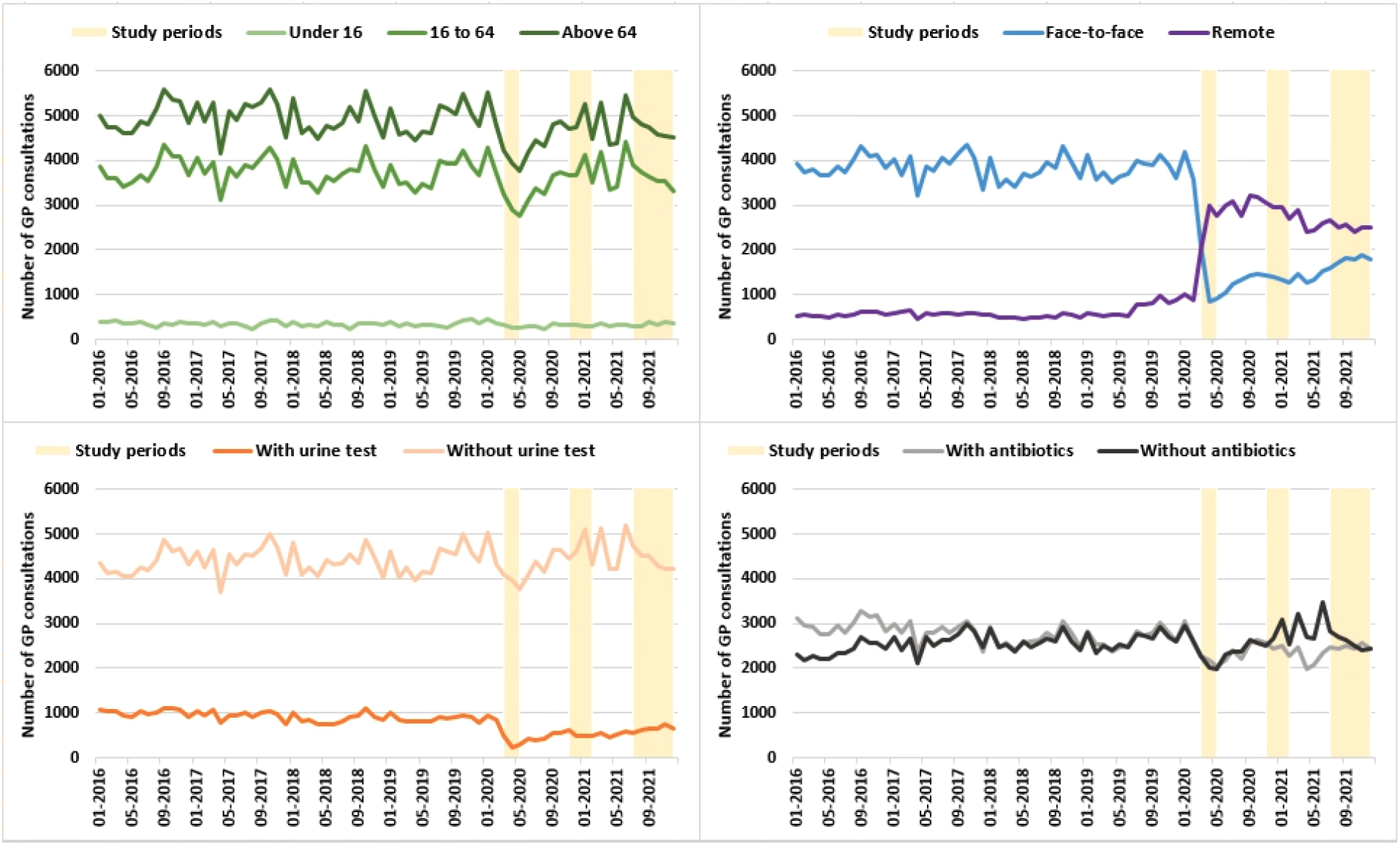
Number of urinary tract infection (UTI) General Practice (GP) consultations per month, by a) age group, b) consultation method, c) diagnosis, and d) with or without antibiotic treatment

### Antibiotic treatment and outcomes of urinary tract infection (UTI): before and during the COVID-19 pandemic

We present the characteristics of GP consultations, antibiotic treatment, and 60-day outcomes based on when the UTI episode was identified (Table 2). Before the pandemic, 72.0% (n = 192,096) of the infections were diagnosed via face-to-face consultation, and 11.2% (n = 29,897) were diagnosed remotely. This has changed to 29.4% (n = 31,977) diagnosed face-to-face and 55.2% (n = 60,146) diagnosed remotely since COVID-19 (Figure 3(b)). The Before the pandemic, 27.2% (n = 4,684) infections in patients under 16 years of age had a diagnosis as per NICE guideline for young patients (i.e., dipstick test, urine culture, or urinalysis) [2], and 16.6% (n = 41,577) infections in patients aged 16 and above had a diagnosis as per NICE guideline for this age group (i.e., urine culture and/or urinalysis) [2]. Since COVID-19, the UTI with diagnosis decreased to 21.1% (n = 1,399) in young patients and 9.7% (n = 9,926) in patients aged 16 and above (Figure 3(c)). In total, we identified 237,495 antibiotic prescriptions (excluding antimycobacterial and anti-leprotic medications) associated with a UTI diagnosis. 52.0% (n = 138,944) of the UTI episodes were treated with antibiotics in the pre-pandemic period, while 47.8% (n = 52,049) were treated during COVID-19 (Figure 3(d)).

**Table 2.** Characteristics, treatment, and outcomes of urinary tract infection (UTI) General Practice (GP) consultations

|  | Pre-pandemic period<br>(Jan 2016 – Feb 2020) |  | Pandemic period<br>(Mar 2020 – Dec 2021) |  | Total |  |
| --- | --- | --- | --- | --- | --- | --- |
|  | N = 266,946 |  | N = 108,913 |  | N = 375,859 |  |
|  | n | % | n | % | n | % |
| **Consultation method** |  |  |  |  |  |  |
| Face-to-face | 192,096 | 72.0% | 31,977 | 29.4% | 224,073 | 59.6% |
| Remote | 29,897 | 11.2% | 60,146 | 55.2% | 90,043 | 24.0% |
| Unknown | 44,953 | 16.8% | 16,790 | 15.4% | 61,743 | 16.4% |
| **Documentation of diagnostic test as per NICE guidelines** |  |  |  |  |  |  |
| In line | 46,261 | 17.3% | 11,325 | 10.4% | 57,586 | 15.3% |
| In line, aged under 16 | 4,684 | 27.2% | 1,399 | 21.1% | 6,083 | 25.5% |
| In line, aged 16 and above | 41,577 | 16.6% | 9,926 | 9.7% | 5,1503 | 14.6% |
| Not in line | 220,685 | 82.7% | 97,588 | 89.6% | 318,273 | 84.7% |
| **Antibiotic treatment** |  |  |  |  |  |  |
| Treated with antibiotics | 138,944 | 52.0% | 52,049 | 47.8% | 190,993 | 50.8% |
| Not treated with antibiotics | 128,002 | 48.0% | 56,864 | 52.2% | 184,866 | 49.2% |
| **Outcome,** |  |  |  |  |  |  |
| 60-day bacteraemia or sepsis | 1,425 | 0.5% | 532 | 0.5% | 1,957 | 0.5% |
| 60-day all-cause mortality | 1,006 | 0.4% | 581 | 0.5% | 1,587 | 0.4% |

Compared to before COVID-19, there has been a decrease in the likelihood of being treated with antibiotics for UTI, most significantly between March and June 2021 (between COVID-19 wave 2 and 3). However, this decrease became insignificant when adjusted for diagnostic testing (i.e., the likelihood of being treated is determined by whether a test was available, and less tests were performed during the pandemic). Regardless of whether the consultation happened before or during the pandemic, antibiotics were more likely to be prescribed if the patients were younger, from non-white ethnic groups, more deprived, with overall better health, had no recent hospital admission, and pregnant (P = 0.000) (Supplementary Material Table S6). Patients who lived in a care home were more likely to be treated with antibiotics (OR = 1.2, p < 0.001, 95% CI = 1.1 to 1.3) compared to those at the same age who did not live in care homes. Having a diagnostic test in line with the guidelines is also a strong predictor for prescribing antibiotic treatment (not tested: OR = 1.0 (reference), tested: OR = 3.2, P = 0.000, 95% CI = 3.2 to 3.3). Whether the initial UTI consultation was delivered via face-to-face or remote methods had no impact on how likely antibiotics were prescribed (P = 0.351).

The most prescribed agents (above 1.0% of the total number of prescriptions) were nitrofurantoin (61.8%, n = 146,485), trimethoprim (20.6%, n = 48,758), amoxicillin (6.2%, n = 14,592), cefalexin (5.6%, n = 13,324), and amoxicillin and clavulanic acid (2.4%, n = 5,709). Course duration was available for 98.8% (n = 234,558) of the UTI antibiotic prescriptions. We present the durations of non-prophylactic, acute prescriptions of each UTI antibiotic agents (nitrofurantoin, trimethoprim, amoxicillin, and cefalexin) in Figure 4. During the pre-pandemic period, 58.7% of the non-prophylactic acute antibiotic prescriptions had durations that exceeded the recommendations, while 49.4% prescriptions exceeded the recommended duration since COVID-19 (Supplementary Material Table S7).

**Figure 4.**
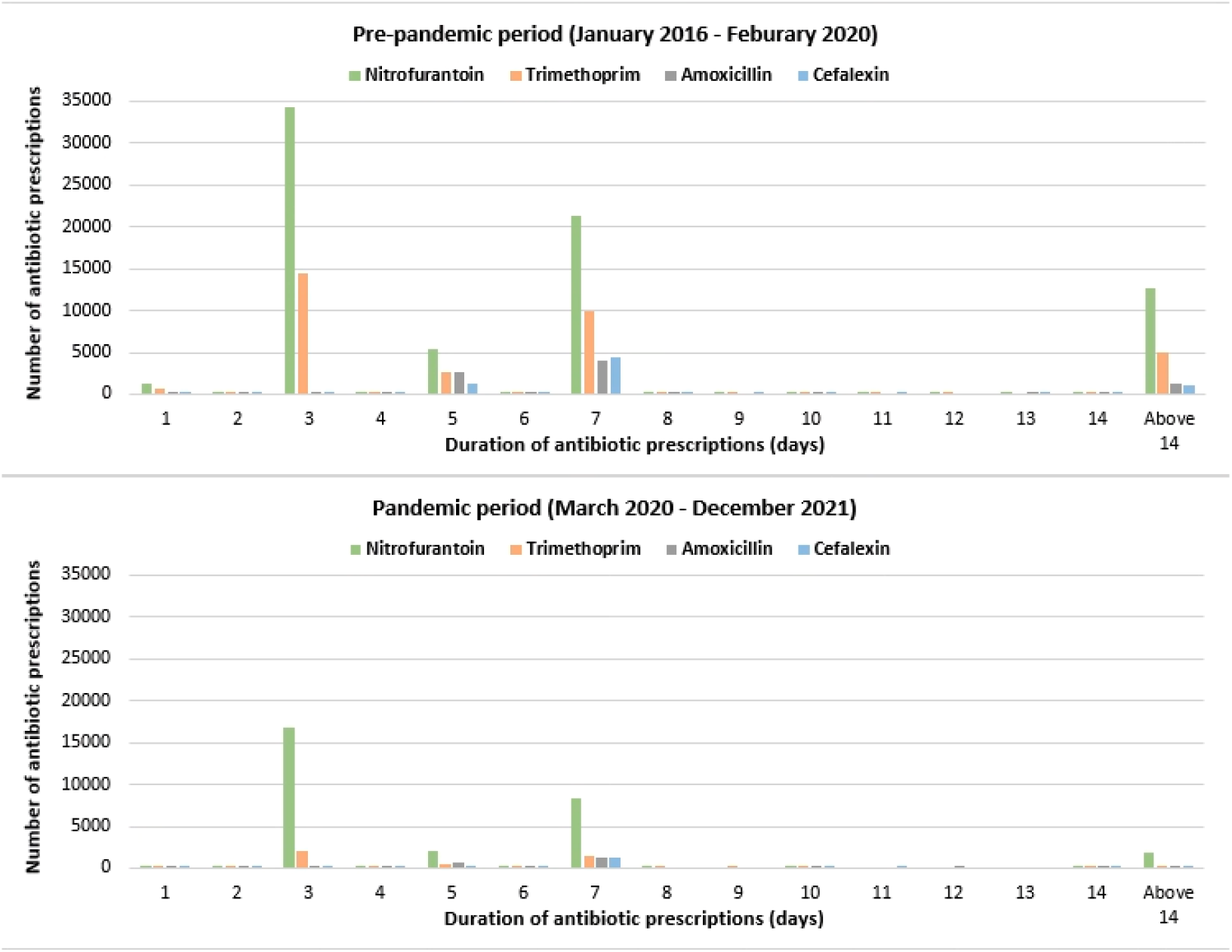
Durations of non-prophylactic, acute antibiotic prescriptions following urinary tract infection (UTI) General Practice (GP) consultations

During the 6-year study period, a total of 1,312,102 days of non-prophylactic acute antibiotic treatment for UTI (nitrofurantoin, trimethoprim, amoxicillin, and cefalexin) were prescribed, of which, 63.3% (830,522 days) were the excess duration beyond the guideline recommendation. Having a diagnostic test is associated with 0.6 fewer days of excess course duration. The excess antibiotic days to treat UTI declined consistently before the COVID-19 pandemic. However, such reduction decelerated since the pandemic onset (p = 0.000) (Figure 5).

**Figure 5.**
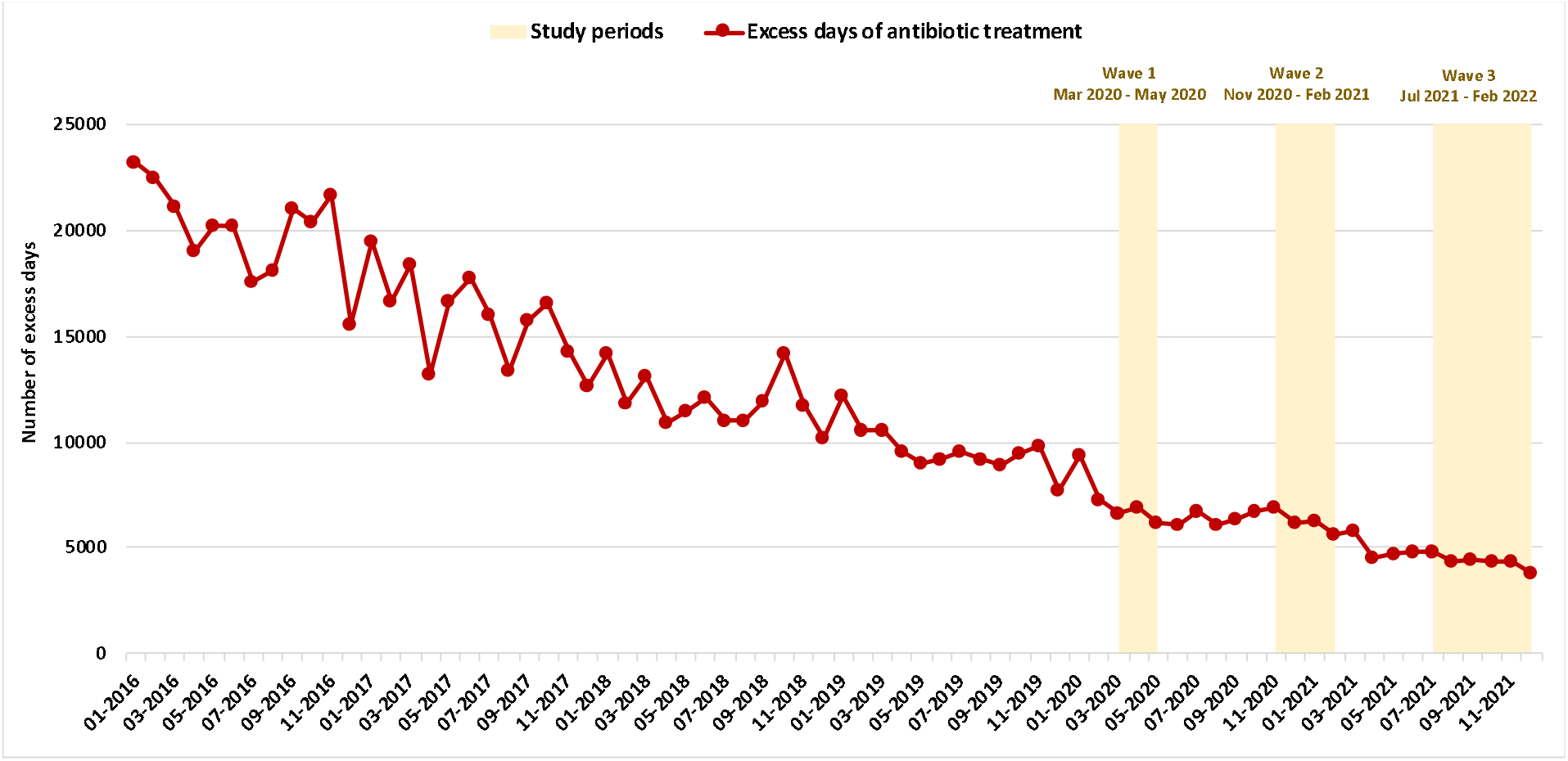
Excess days of antibiotic treatment for urinary tract infection (UTI)

Overall, 0.5% (n = 1,957) of the UTI patients had bacteraemia or sepsis that required GP re-consultation or/and hospital admission within 60 days following the initial UTI diagnosis. The percentage of UTI associated with subsequent bacteraemia or sepsis within the next 60 days significantly increased among patients who were not treated with antibiotics (without antibiotics: 0.9%, with antibiotics: 0.2%, p = 0.000). After adjusting for age, ethnicity, level of deprivation, co-morbidities, recent hospitalisation, and diagnostic testing, patients who received no antibiotic treatment were significantly more likely to experience bacteraemia or sepsis within 60 days compared with those who were treated with antibiotics (without antibiotics: aOR = 1.0 (reference), with antibiotics: aOR = 0.1, P = 0.008, 95% CI = 0.0 to 0.6). The risk of having bacteraemia or sepsis in the next 60 days following a UTI did not change significantly during the pandemic (Supplementary Material Table S7). Temporally, a higher proportion of UTI episodes that had subsequent bacteraemia or sepsis correlated with lower prescribing level during COVID-19 wave 2 and 3 (Figure 3d; Supplementary Material Figure S1), however, such increase was not significant. During wave 3, the proportion of UTI episodes that progressed to bacteraemia or sepsis decreased significantly (Supplementary Material Table S8, Figure S1). The percentage of UTI with 60-day all-cause mortality was 0.4% (n = 1,587) across the study period among all patients, 0.5% without antibiotics, and 0.4% with antibiotics, after excluding those who had laboratory-confirmed SARS-CoV-2 infection. The adjusted odds ratio of experiencing mortality within 60 days is 0.8 for infections treated by antibiotics (P = 0.000, 95% CI = 0.8 to 0.9).

## Discussion

In this analysis, we examined epidemiological patterns, management, and outcomes of community-onset UTI in female patients in North West London before and during the COVID-19 pandemic. During the pre-pandemic years, UTI GP consultation rates stayed level with seasonal peaking in October every year. We observed declines in UTI GP consultation rate coincident with surges in COVID-19 cases. Steep upward shifts in UTI GP consultation rate occurred when COVID-19 cases fell. Multiple factors may have driven the observed infection patterns in UTI. This includes changes in health seeking behaviours during COVID-19, such that that many patients who experienced UTI symptoms voluntarily chose to avoid visiting healthcare facilities, due to fear of COVID-19 infection or desire to avoid placing additional pressure on the health system. Difficulty in access to primary care service when local health systems reached the maximum capacity is also likely to have had impact on UTI presentations [21,22]. Additionally, improved environmental and personal hygiene, as a result of COVID-19 measures, may have potentially reduced the occurrence of UTIs in the community.

In April 2020, a rapid government mandated shift to remote primary care consultation took place, which allowed focus on vulnerable patients who were older, shielding, and with poorer overall health [9,23,24]. The percentage of GP consultations delivered face-to-face decreased significantly since COVID-19, from a pre-pandemic average of 72.0% to 22.6% in May 2020. Less UTI diagnoses were made with urine culture and urinalysis during the pandemic. The UTI consultations supported by appropriate diagnostic testing, in line with the NICE guidelines, were more likely to be followed by antibiotic treatment with the recommended course duration. Nitrofurantoin, trimethoprim, amoxicillin, and cefalexin were commonly prescribed antibiotics following the UTI consultation before and during the pandemic. The absence of antibiotic treatment is associated with a 10-fold increase in the risk of experiencing bacteraemia or sepsis, consistent with the previous studies conducted among patients aged older than 65 years (both female and male) across England [3,5]. Though the patients who consulted GP during the pandemic were older and more co-morbid, and remote consultation and less testing increased uncertainty in diagnosis, there has been no significantly increased risk of having bacteraemia or sepsis following UTI since COVID-19. While not receiving antibiotics after a UTI consultation was associated with higher risk of developing bacteraemia or sepsis, using antibiotics with unnecessarily long duration is also concerning as the latter drives the emergence of AMR. Durations of recommended antibiotic treatment for UTI showed poor guideline adherence. Between January 2016 and December 2021, our study cohort received 830,522 excess days of non-prophylactic acute antibiotic treatment for UTI (nitrofurantoin, trimethoprim, amoxicillin, cefalexin). Before the COVID-19 pandemic, the excess days of antibiotic treatment for UTI had achieved a consistent reduction, demonstrating the positive impact of national stewardship interventions to improve UTI treatment, such as Quality Premium (QP) [25–27]. In our analysis, we found that the antibiotic treatment prescribed for the UTI cases supported by diagnostic testing were less likely to exceed the guideline recommendation. This might explain why the decline in excess antibiotic days slowed down since COVID-19, as the general practitioners had the tendency to choose longer courses when prescribing remotely, having less confidence in the assessment.

Overall, we observed a low level of diagnostic testing to support UTI consultations before and during the pandemic period, which leads to the question of whether antibiotics were prescribed to treat self-limiting urinary tract symptoms. More specific guidelines are required to better advise patients to manage sometimes painful urinary tract symptoms without antibiotics. In addition, back-up antibiotic prescription (to use if symptoms do not start to improve within 48 hours or worsen at any time) has been recommended as a national strategy [2,28] to reduce unnecessary antibiotic use while ensuring patients can access antibiotics when needed. In practice, GP prescribers often do not indicate whether an antibiotic prescription was a back-up option electronically, despite standard codes provided to support documentation [29,30]. This brings additional challenge to evaluation of the utilisation of back-up prescribing and the impact on patient outcomes, therefore this issue should be addressed when further studies are conducted. In previous studies, researchers defined antibiotic prescribed on the same day of the UTI consultation “immediate”, and the ones prescribed between 2 to 7 days after the UTI consultation “delayed” or “deferred” [3,5]. However, such definitions do not reflect how back-up prescriptions are used in real practice. For instance, antibiotics may often be prescribed on the same day of the UTI consultation, with the patient instructed to “wait and see” [31]. Such prescriptions would be incorrectly defined as “immediate”. Improving utilisation of standard primary care coding for prescriptions as well as procedures and observations is urgently needed to allow accurate and robust assessment of such national strategies.

Our study has three main limitations. First, we did not include microbiology laboratory data to ascertain the causative pathogens and resistance profiles. We overcame this by triangulating the trends of UTI GP consultation rate derived using GP diagnosis with what has been reported at regional level based on aggregated microbiology data [32]. Second, we cannot separate community-onset and hospital-onset bacteraemia, especially during the COVID pandemic, due to the reduced hospital admission during COVID, which may lead of an over-estimation of hospital onset cases (i.e., patient had shorter LOS and therefore reduced the denominator patient-days) despite the steep decline in case numbers. The third limitation is the potential over-representation / over-estimation of face-to-face appointments due to inaccurate documentation of consultation method. This limitation highlighted, again, the importance to improve the quality of clinical coding in primary care.

To conclude, our study provides a comprehensive examination of management and outcomes of community-onset UTI in female patients, considering the changes in GP consultations during the COVID-19 pandemic. Continued monitoring is required to assess the overall impact on patients and health systems from the changed landscape of primary care delivery.

## Data Availability

All data produced in the present study are available upon reasonable request to the authors

## Notes

### Author Contributions

NZ, AH and PA developed the concept and methodology for this research. NZ undertook data collection and analysis. NZ, BH and RA drafted the initial manuscript. RA, JRP, DL, MM, EF, TMR, PA and AH contributed significantly to data interpretation, revision of the manuscript and finalisation for submission. PA is the guarantor of the study. The corresponding author attests that all listed authors meet the ICMJE criteria for authorship and that no other meeting the criteria have been omitted.

### Transparency statement

The lead author affirms that the manuscript is an honest, accurate, and transparent account of the study being reported; that no important aspects of the study have been omitted; and that any discrepancies from the study as originally planned (and, if relevant, registered) have been explained.

### Disclaimer

The views expressed in this publication are those of the author(s) and not necessarily those of the NHS, the National Institute for Health Research, the Department of Health and Social Care, or UK Health Security Agency.

### Financial support and acknowledgement

This research was funded by 1) the World Health Organization (WHO), 2) the National Institute for Health Research (NIHR) Health Protection Research Unit (HPRU) in Healthcare Associated Infections and Antimicrobial Resistance (HCAI & AMR) at Imperial College London in partnership with the UK Health Security Agency (previously Public Health England (PHE)), in collaboration with, Imperial Healthcare Partners, University of Cambridge and University of Warwick, and 3) the Department of Health and Social Care, who funded Centre for Antimicrobial Optimisation (CAMO) at Imperial College London. AH is a NIHR Senior Investigator. PA is supported by the NIHR Applied Research Collaboration Northwest London. RA acknowledges support from Wellcome Trust as the deputy chair of the Surveillance and Epidemiology of Drug-resistant Infections Consortium (SEDRIC). BH is grateful for the support of the NIHR under the Applied Health Research (ARC) programme for North West London. This report is independent research funded by the National Institute for Health Research. The views expressed in this publication are those of the author(s) and not necessarily those of the WHO, the National Institute for Health Research, the Department of Health and Social Care or the UK Health Security Agency.

