## Supplementary Material for "Community-onset urinary tract infection in females in the context of COVID-19: a longitudinal population cohort study exploring case presentation, management, and outcomes"

**Table S1. Suspected UTI GP diagnosis in READ and Systematized Nomenclature of Medicine Clinical Terms (SNOMED-CT)**

| **READ** | **Version** | **READ diagnosis** | **Core concept ID** | **SNOMED-CT** | **SNOMED-CT diagnosis** |
| --- | --- | --- | --- | --- | --- |
| 1J4..00 | 2 | Suspected UTI | 210843 | 314940005 | Suspected urinary tract infection (situation) |
| 68566 | 3 | Urinary tract infection |  |  |  |
|  |  |  | 267015 | 431309003 | Acute urinary tract infection |
| K190. | 2 | Urinary tract infection, unspecified site | 57494 | 68566005 | Urinary tract infectious disease (disorder) |
| K190. | 3 | Urinary tract infection: [site not specified] or [recurrent] |  |  |  |
| K1900 | 2 | Bacteriuria, site not specified | 51469 | 61373006 | Bacteriuria (finding) |
| K1900 | 3 | Bacteriuria: [site not specified] or [asymptomatic] | 311296 | 720406004 | Asymptotic bacteriuria (finding) |
| K1900-1 | 2 | Asymptotic bacteriuria | 311296 | 720406004 | Asymptotic bacteriuria (finding) |
| K1901 | 2 | Pyuria, site not specified | 4003 | 4800001 | Pyuria (finding) |
| K1901 | 3 | Pyuria | 4003 | 4800001 | Pyuria (finding) |
| K190-1 | 2 | Recurrent urinary tract infection | 124325 | 197927001 | Recurrent urinary tract infection (disorder) |
| K190-98 | 2 | Urinary tract infection NOS | 267015 | 431309003 | Acute urinary tract infection |
| K190-99 | 2 | Urine infection | 267015 | 431309003 | Acute urinary tract infection |
| K1902 | 2 | Post operative UTI | 124324 | 197926005 | Postoperative urinary tract infection (disorder) |
| K1902 | 3 | Postoperative urinary tract infection | 124324 | 197926005 | Postoperative urinary tract infection (disorder) |
| K1903 | 2 | Recurrent urinary tract infection | 124325 | 197927001 | Recurrent urinary tract infection (disorder) |
| K1903 | 3 | Recurrent urinary tract infection | 124325 | 197927001 | Recurrent urinary tract infection (disorder) |
| K1903-1 | 2 | Recurrent UTI | 124325 | 197927001 | Recurrent urinary tract infection (disorder) |
| K1904 | 2 | Chronic urinary tract infection | 124326 | 197928006 | Chronic urinary tract infection (disorder) |
| K1904 | 3 | Chronic urinary tract infection | 124326 | 197928006 | Chronic urinary tract infection (disorder) |
| K1905 | 2 | Urinary tract infection | 57494 | 68566005 | Urinary tract infectious disease (disorder) |
| K1906 | 2 | Urosepsis | 311838 | 721104000 | Sepsis due to urinary tract infection |
| K190x | 2 | Persist proteinuria, unspec | 335591 | 12511000132108 | Persist proteinuria (finding) |
| K190z | 2 | Urinary tract infection, site unspecified NOS | 57494 | 68566005 | Urinary tract infectious disease (disorder) |
| K190z | 3 | Urinary tract infection, site not specified NOS | 57494 | 68566005 | Urinary tract infectious disease (disorder) |
| K190z00 |  | Urinary tract infection, site not specified NOS | 2821458 | 197930008 | Urinary tract infection, site not specified NOS |
|  |  |  | 57494 | 155897002 | Urinary tract infection (&[NOS]) |
|  |  |  | 57494 | 266635000 | Urinary tract infection (&[NOS]) |
|  |  |  | 57494 | 197924008 | Urinary tract infection [site not specified] or [recurrent] |
| K15.. | 2 | Cystitis | 32515 | 38822007 | Cystitis |
| K15.. | 3 | Inflammation of bladder | 32515 | 38822007 | Cystitis |
| K15..00 | 3 | Cystitis | 32515 | 38822007 | Cystitis |
| K150. | 2 | Acute cystitis | 57203 | 68226007 | Acute cystitis |
| K15y. | 2 | Other specified cystitis | 32515 | 38822007 | Cystitis |
| K15y. | 3 | Other specified cystitis | 32515 | 38822007 | Cystitis |
| K15yz | 2 | Other cystitis NOS | 32515 | 38822007 | Cystitis |
| K15yz | 3 | Other cystitis NOS | 32515 | 38822007 | Cystitis |
| K15z. | 2 | Cystitis NOS | 32515 | 38822007 | Cystitis |
| K15z. | 3 | Cystitis NOS | 32515 | 38822007 | Cystitis |
| K15z.00 | 3 | Cystitis NOS | 2610839 | 197857009 | Cystitis NOS |
| K152. | 2 | Other chronic cystitis | 28154 | 33655002 | Chronic cystitis (disorder) |
| K152. | 3 | Other chronic cystitis | 28154 | 33655002 | Chronic cystitis (disorder) |
| K1520 | 2 | Subacute cystitis | 3597 | 4324001 | Subacute cystitis (disorder) |
| K1520 | 3 | Subacute cystitis | 3597 | 4324001 | Subacute cystitis (disorder) |
| K152y | 2 | Chronic cystitis unspecified | 3597 | 33655002 | Subacute cystitis (disorder) |
| K152y | 3 | Chronic cystitis unspecified | 3597 | 33655002 | Subacute cystitis (disorder) |
| K152y-99 | 2 | Chronic cystitis |  | 33655002 |  |
| K152z | 2 | Other chronic cystitis NOS | 28154 | 33655002 | Chronic cystitis (disorder) |
| K152z | 3 | Other chronic cystitis NOS | 28154 | 33655002 | Chronic cystitis (disorder) |
|  |  |  | 145350 | 236702008 | Acute culture positive cystitis |
|  |  |  | 145281 | 236624004 | Acute culture negative cystitis |
| K155. | 2 | Recurrent cystitis | 124305 | 197853008 | Recurrent cystitis |
| K155. | 3 | Recurrent cystitis | 124305 | 197853008 | Recurrent cystitis |
| Kyu50 | 2 | [X]Other chronic cystitis | 28154 | 33655002 | Chronic cystitis (disorder) |
| Kyu50 | 3 | [X]Other chronic cystitis | 28154 | 33655002 | Chronic cystitis (disorder) |
| Kyu51 | 2 | [X]Other cystitis | 32515 | 38822007 | Cystitis |
| Kyu51 | 3 | [X]Other cystitis | 32515 | 38822007 | Cystitis |
| 1AG.. | 2 | Recurrent urinary tract infection | 124325 | 197927001 | Recurrent urinary tract infection (disorder) |
| 1AZ6. | 2 | Lower urinary tract symptoms | 205308 | 307541003 | Lower urinary tract symptoms (finding) |
| XaB9O | 3 | Lower urinary tract symptoms | 205308 | 307541003 | Lower urinary tract symptoms (finding) |
| 1AZ60 | 2 | Mild low urinary tract symptoms | 353122 | 763121000000102 | Mild low urinary tract symptoms (finding) |
| XaXHi | 3 | Mild low urinary tract symptoms | 353122 | 763121000000102 | Mild low urinary tract symptoms (finding) |
| 1AZ61 | 2 | Moderate lower urinary tract symptoms | 353120 | 763121000000108 | Moderate lower urinary tract symptoms (finding) |
| XaXHj | 3 | Moderate lower urinary tract symptoms | 353120 | 763121000000108 | Moderate lower urinary tract symptoms (finding) |
| 1AZ62 | 2 | Severe lower urinary tract symptoms | 353119 | 763121000000106 | Severe lower urinary tract symptoms (finding) |
| XaXHk | 3 | Severe lower urinary tract symptoms | 353119 | 763121000000106 | Severe lower urinary tract symptoms (finding) |
| Xa8EJ | 3 | Acute cystitis | 57203 | 68226007 | Acute cystitis |
|  |  |  | 205308 | 706721000000105 | Lower urinary tract symptoms (finding) |
| SP077 | 2 | Inf+infl r/pr dev, implt+g ur s | 130118 | 213137004 | Infection inflammation reaction due to prosthetic device, implant and graft in urinary system (disorder) |
| SP07Q | 2 | Catheter-associated UTI | 294613 | 700372006 | Urinary tract infection associated with catheter (disorder) |
| XaaZd | 3 | Catheter-associated urinary tract infection | 294613 | 700372006 | Urinary tract infection associated with catheter (disorder) |
| X30PX | 3 | Lower urinary tract infection | 3332 | 4009004 | Lower urinary tract infectious disease (disorder) |
| XE0e0 | 3 | Infection of urinary tract | 57494 | 68566005 | Urinary tract infectious disease (disorder) |
| XE0e1 | 3 | Bacteriuria | 51469 | 61373006 | Bacteriuria (finding) |
| XE0fj | 3 | Urinary tract infection (&[NOS]) | 57494 | 68566005 | Urinary tract infectious disease (disorder) |
| XE0fl | 3 | (urinary obstruction unsp) or (recur urinary tract infection) | 101402 | 128606002 | Disorder of the urinary system (disorder) |
| X30Pv | 3 | Symptomatic disorders of the urinary tract | 145360 | 236714000 | Symptomatic disorders of the urinary tract |
| 14D4. | 2 | H/O: recurrent cystitis | 108640 | 161549001 | History of recurrent cystitis (situation) |
| 14D4. | 3 | H/O: recurrent cystitis | 108640 | 161549001 | History of recurrent cystitis (situation) |
| 14D7. | 2 | H/O: recurrent UTI | 291091 | 473116008 | History of recurrent urinary tract infection (situation) |
| 1428 | 2 | H/O: * urinary system | 251984 | 197853008 | History of urinary system |
| 1428 | 3 | H/O: * urinary system (&[bladder] or [kidney]) |  |  |  |
| X30Na | 3 | Recurrent cystitis (culture negative | 145282 | 236625003 | Recurrent cystitis (culture negative) (disorder) |
| X30Nb | 3 | Chronic cystitis (culture negative | 145283 | 236625002 | Chronic cystitis (culture negative) (disorder) |
| X30Nc | 3 | Chronic nonspecific cystitis | 145284 | 236627006 | Chronic nonspecific cystitis (disorder) |
| 7N51. | 2 | Lower urinary tract | 16487 | 19787009 | Lower urinary tract structure (body structure) |
| 7N51. | 3 | [SO]Lower urinary tract | 16487 | 19787009 | Lower urinary tract structure (body structure) |
| Xaglc | 3 | Uncomplicated urinary tract infection | 373676 | 1090711000000100 | Uncomplicated urinary tract infection (disorder) |
| L1666 | 2 | Urinary tract infection following delivery | 124642 | 199111004 | Urinary tract infection following delivery (disorder) |
| L1666 | 3 | Urinary tract infection following delivery | 124642 | 199111004 | Urinary tract infection following delivery (disorder) |
| L1668 | 2 | UTI complicating pregnancy | 205306 | 307534009 | Urinary tract infection in pregnancy |
| L166z | 3 | UTI complicating pregnancy (& [NOS]) | 205306 | 307534009 | Urinary tract infection in pregnancy |
| L166z-1 | 2 | UTI - urinary tract infection in pregnancy | 205306 | 307534009 | Urinary tract infection in pregnancy |
| L166z-1 |  |  | 2630166 | 199115008 | UTI in pregnancy |
| L1668 |  |  | 2617501 | 199114007 | Urinary tract infection complicating pregnancy |
| L1668 |  |  | 2593802 | 309782006 | Urinary tract infection complicating pregnancy |
| Xa7nb | 3 | Coliform urinary tract infection | 199591 | 301010001 | Coliform urinary tract infection (disorder) |
| Xa7nc | 3 | Escherichia coli urinary tract infection | 199592 | 301011002 | Escherichia coli urinary tract infection (disorder) |
| Xa7ne | 3 | Pseudomonas urinary tract infection | 199593 | 301011004 | Pseudomonas urinary tract infection (disorder) |
|  |  |  | 346201 | 368991000119100 | Urinary tract infection caused by Enterococcus |
|  |  |  | 346202 | 369001000119100 | Urinary tract infection caused by Klebsiella |
|  |  |  | 346203 | 369001000119102 | Urinary tract infection caused by Pseudomonas |

**Table S2. Bacteraemia, bloodstream infection, or sepsis GP diagnosis in READ and Systematized Nomenclature of Medicine Clinical Terms (SNOMED-CT)**

| **READ** | **Version** | **READ diagnosis** | **Core concept ID** | **SNOMED-CT**  **(Scheme 71)** | **SNOMED-CT diagnosis** |
| --- | --- | --- | --- | --- | --- |
| A2701 | 2 | Listeria septicaemia | 281800 | 449335002 | Sepsis caused by Listeria monocytogenes (disorder) |
| A2701 | 3 | Listeria septicaemia | 281800 | 449335002 | Sepsis caused by Listeria monocytogenes (disorder) |
| A2706 | 2 | Sepsis Listeria monocytogenes | 281800 | 449335002 | Sepsis caused by Listeria monocytogenes (disorder) |
| A2711 | 2 | Erysipelothrix septicaemia | 280167 | 447684006 | Sepsis caused by Erysipelothrix (disorder) |
| A2711 | 3 | Erysipelothrix septicaemia | 280167 | 447684006 | Sepsis caused by Erysipelothrix (disorder) |
| A2713 | 3 | Sepsis due to Erysipelothrix | 280167 | 447684006 | Sepsis caused by Erysipelothrix (disorder) |
| A38.. | 2 | Septicaemia | 76601 | 91302008 | Sepsis (disorder) |
| A38.. | 3 | Septicaemia | 76601 | 91302008 | Sepsis (disorder) |
| A380. | 2 | Streptococcal septicaemia | 280896 | 448418006 | Sepsis caused by Streptococcus (disorder) |
| A380. | 3 | Streptococcal septicaemia | 280896 | 448418006 | Sepsis caused by Streptococcus (disorder) |
| A3800 | 2 | Septicaem due streptococc gp A | 281966 | 449504009 | Sepsis caused by Streptococcus pyogenes (disorder) |
| A3800 | 3 | Gp A streptococcal septicaemia | 281966 | 449504009 | Sepsis caused by Streptococcus pyogenes (disorder) |
| A3801 | 2 | Septicaem due streptococc gp B | 280897 | 448419003 | Sepsis caused by Streptococcus agalactiae (disorder) |
| A3801 | 3 | Gp B streptococcal septicaemia | 280897 | 448419003 | Sepsis caused by Streptococcus agalactiae (disorder) |
| A3802 | 2 | Septicaem due streptococc gp D | 280898 | 448420009 | Sepsis caused by Streptococcus group D (disorder) |
| A3802 | 3 | Gp D streptococcal septicaemia | 280898 | 448420009 | Sepsis caused by Streptococcus group D (disorder) |
| A3803 | 2 | Septicaem due strep pneumon | 280899 | 448421008 | Sepsis caused by Streptococcus pneumoniae (disorder) |
| A3804 | 2 | Septicaem due enterococcus | 207796 | 310669007 | Sepsis caused by enterococcus (disorder) |
| A3805 | 2 | Vancomy resist enteroc septic | 207778 | 310649002 | Vancomycin resistant enterococcal septicaemia (disorder) |
| A381. | 2 | Staphylococcal septicaemia | 280376 | 447894003 | Sepsis caused by Staphylococcus (disorder) |
| A381. | 3 | Staphylococcal septicaemia | 280376 | 447894003 | Sepsis caused by Staphylococcus (disorder) |
| A3810 | 2 | Septicaem due to staph aureus | 280895 | 448417001 | Sepsis caused by Staphylococcus aureus (disorder) |
| A3810 | 3 | Septicaemia due to Staphylococcus aureus | 280895 | 448417001 | Sepsis caused by Staphylococcus aureus (disorder) |
| A3811 | 2 | Septicaem, coag-neg staphylococ | 281967 | 449505005 | Sepsis caused by coagulase-negative Staphylococcus (disorder) |
| A3811 | 3 | Septicaemia due to coagulase-negative staphylococcus | 281967 | 449505005 | Sepsis caused by coagulase-negative Staphylococcus (disorder) |
| A382. | 2 | Pneumococcal septicaemia | 280899 | 448421008 | Sepsis caused by Streptococcus pneumoniae (disorder) |
| A382. | 3 | Pneumococcal septicaemia | 280899 | 448421008 | Sepsis caused by Streptococcus pneumoniae (disorder) |
| A383. | 2 | Septicaemia due to anaerobes | 280325 | 447843005 | Sepsis caused by anaerobic bacteria (disorder) |
| A383. | 3 | Septicaemia due to anaerobes | 280325 | 447843005 | Sepsis caused by anaerobic bacteria (disorder) |
| A384. | 2 | Septicaemia due to other Gram-negative organisms | 281550 | 449082003 | Sepsis caused by Gram negative bacteria (disorder) |
| A384. | 3 | Septicaemia – other gram -ve | 281550 | 449082003 | Sepsis caused by Gram negative bacteria (disorder) |
| A3840 | 2 | Gram-negative septicaemia NOS | 281550 | 449082003 | Sepsis caused by Gram negative bacteria (disorder) |
| A3840 | 3 | Gram-negative septicaemia NOS | 281550 | 449082003 | Sepsis caused by Gram negative bacteria (disorder) |
| A3841 | 2 | Haemophilus infl. septicaemia | 280168 | 447685007 | Sepsis caused by Haemophilus influenzae (disorder) |
| A3841 | 3 | Haemophilus influenza septicaemia | 280168 | 447685007 | Sepsis caused by Haemophilus influenzae (disorder) |
| A3842 | 2 | Escherichia coli septicaemia | 280381 | 447899008 | Sepsis caused by Escherichia coli (disorder) |
| A3842 | 3 | Escherichia coli septicaemia | 280381 | 447899008 | Sepsis caused by Escherichia coli (disorder) |
| A3843 | 2 | Pseudomonas septicaemia | 281287 | 448813005 | Sepsis caused by Pseudomonas (disorder) |
| A3843 | 3 | Pseudomonas septicaemia | 281287 | 448813005 | Sepsis caused by Pseudomonas (disorder) |
| A3844 | 2 | Serratia septicaemia | 281552 | 449084002 | Sepsis caused by Serratia (disorder) |
| A3844 | 3 | Serratia septicaemia | 281552 | 449084002 | Sepsis caused by Serratia (disorder) |
| A384z | 2 | Other gram-ve septicaemia NOS | 281550 | 449082003 | Sepsis caused by Gram negative bacteria (disorder) |
| A384z | 3 | Other gram-negative septicaemia NOS | 281550 | 449082003 | Sepsis caused by Gram negative bacteria (disorder) |
| A38y. | 2 | Other specified septicaemias | 76601 | 91302008 | Sepsis (disorder) |
| A38y. | 3 | Other specified septicaemias | 76601 | 91302008 | Sepsis (disorder) |
| A38z. | 2 | (Septicaemia NOS) or (sepsis) |  |  |  |
| A38z. | 3 | Septicaemia NOS | 76601 | 91302008 | Sepsis (disorder) |
| A395. | 2 | Actinomycotic septicaemia | 280380 | 447898000 | Sepsis caused by Actinomyces (disorder) |
| A395. | 3 | Actinomycotic septicaemia | 280380 | 447898000 | Sepsis caused by Actinomyces (disorder) |
| A396.00 | 2 | Sepsis due to Actinomyces | 280380 | 447898000 | Sepsis caused by Actinomyces (disorder) |
| A3C.. | 2 | Sepsis | 76601 | 91302008 | Sepsis (disorder) |
| A3C0. | 2 | Sepsis due to Streptococcus | 280896 | 448418006 | Sepsis caused by Streptococcus (disorder) |
| A3C00 | 2 | Sepsis Streptococcus group A | 281966 | 449504009 | Sepsis caused by Streptococcus pyogenes (disorder) |
| A3C01 | 2 | Sepsis Streptococcus group B | 280897 | 448419003 | Sepsis caused by Streptococcus agalactiae (disorder) |
| A3C02 | 2 | Sepsis Streptococcus group D | 280897 | 448419003 | Sepsis caused by Streptococcus group D (disorder) |
| A3C03 | 2 | Sepsis Streptococcus pneumonia | 280897 | 448419003 | Sepsis caused by Streptococcus pneumoniae (disorder) |
| A3C0y | 2 | Other streptococcal sepsis | 280896 | 448418006 | Sepsis caused by Streptococcus (disorder) |
| A3C0z | 2 | Streptococcal sepsis unspecifi | 280896 | 448418006 | Sepsis caused by Streptococcus (disorder) |
| A3C1. | 2 | Sepsis due to Staphylococcus | 280376 | 447894003 | Sepsis caused by Staphylococcus (disorder) |
| A3C10 | 2 | Sepsis Staphylococcus aureus | 280895 | 448417001 | Sepsis caused by Staphylococcus aureus (disorder) |
| A3C1y | 2 | Sepsis oth spec staphylococcus | 280376 | 447894003 | Sepsis caused by Staphylococcus (disorder) |
| A3C1z | 2 | Sepsis due staphylococcus NOS | 280376 | 447894003 | Sepsis caused by Staphylococcus (disorder) |
| A3C2. | 2 | Sepsis due anaerobic bacteria | 280325 | 447843005 | Sepsis caused by anaerobic bacteria (disorder) |
| A3C3. | 2 | Sepsis Gram negative bacteria | 281550 | 449082003 | Sepsis caused by Gram negative bacteria (disorder) |
| A3C30 | 2 | Sepsis Haemophilus influenzae | 280168 | 447685007 | Sepsis caused by Haemophilus influenzae (disorder) |
| A3C3y | 2 | Sepsis oth Gram neg organisms | 281550 | 449082003 | Sepsis caused by Gram negative bacteria (disorder) |
| A3Cy. | 2 | Other specified sepsis | 76601 | 91302008 | Sepsis (disorder) |
| A3Cz. | 2 | Sepsis NOS | 76601 | 91302008 | Sepsis (disorder) |
| AB2y3 | 2 | Candidal septicaemia | 280323 | 447841007 | Sepsis caused by Candida (disorder) |
| AB2y3 | 3 | Candidal septicaemia | 280323 | 447841007 | Sepsis caused by Candida (disorder) |
| AB2y5 | 2 | Candidal sepsis | 280323 | 447841007 | Sepsis caused by Candida (disorder) |
| Ayu3E00 | 2 | [X] Oth streptococl septicaemia | 280896 | 448418006 | Sepsis caused by Streptococcus (disorder) |
| Ayu3E00 | 3 | [X] Other streptococcal septicaemia | 280896 | 448418006 | Sepsis caused by Streptococcus (disorder) |
| Ayu3F00 | 2 | [X] Streptococcal septicaemia, unspecified | 280896 | 448418006 | Sepsis caused by Streptococcus (disorder) |
| Ayu3F00 | 3 | [X] Streptoc septicaemia, unspecified | 280896 | 448418006 | Sepsis caused by Streptococcus (disorder) |
| Ayu3G00 | 2 | [X] Septicaem/oth gram-ve orgnsm | 281550 | 449082003 | Sepsis caused by Gram negative bacteria (disorder) |
| Ayu3G00 | 3 | [X] Septicaemia due to other gram-negative organisms | 281550 | 449082003 | Sepsis caused by Gram negative bacteria (disorder) |
| Ayu3H00 | 2 | [X] Other specified septicaemia | 76601 | 91302008 | Sepsis (disorder) |
| Ayu3H00 | 3 | [X] Other specified septicaemia | 76601 | 91302008 | Sepsis (disorder) |
| Ayu3J00 | 2 | [X] Septicaemia, unspecified | 76601 | 91302008 | Sepsis (disorder) |
| Ayu3J00 | 3 | [X] Septicaemia, unspecified | 76601 | 91302008 | Sepsis (disorder) |
| K1906 | 2 | Urosepsis | 311838 | 721104000 | Sepsis due to urinary tract infection (disorder) |
| R106. | 2 | [D] Unspecified bacteraemia | 4808 | 5758002 | Bacteremia (finding) |
| R106. | 3 | [D] Unspecified bacteraemia | 4808 | 5758002 | Bacteremia (finding) |

**Table S3. Bacteraemia, bloodstream infection, or sepsis hospital diagnosis in International Statistical Classification of Diseases and Health Related Problems version 10 (ICD-10)**

| **ICD-10** | **ICD-10 diagnosis (Scheme 1302012)** | **ICD-10 Concept ID** |
| --- | --- | --- |
| A02.1 | Salmonella sepsis | 1302031 |
| A32.7 | Listerial sepsis | 1302204 |
| A39.2 | Acute meningococcaemia | 1302226 |
| A39.3 | Chronic meningococcaemia | 1302227 |
| A39.4 | Meningococcaemia, unspecified | 1302228 |
| A40 | Streptococcal sepsis | 1302232 |
| A40.0 | Sepsis due to streptococcus, group A | 1302233 |
| A40.1 | Sepsis due to streptococcus, group B | 1302234 |
| A40.2 | Sepsis due to streptococcus, group D | 1302235 |
| A40.3 | Sepsis due to streptococcus pneumoniae | 1302236 |
| A40.8 | Other streptococcal sepsis | 1302237 |
| A40.9 | Streptococcal sepsis, unspecified | 1302238 |
| A41 | Other septicaemia | 1302239 |
| A41.0 | Sepsis due to Staphylococcus aureus | 1302240 |
| A41.1 | Sepsis due to other specified staphylococcus | 1302241 |
| A41.2 | Sepsis due to unspecified staphylococcus | 1302242 |
| A41.3 | Sepsis due to Haemophilus influenzae | 1302243 |
| A41.4 | Sepsis due to anaerobes | 1302244 |
| A41.5 | Sepsis due to other Gram-negative organisms | 1302245 |
| A41.51 | Sepsis due to Escherichia coli [E. coli] |  |
| A41.52 | Sepsis due to Pseudomonas |  |
| A41.53 | Sepsis due to Serratia |  |
| A41.59 | Other Gram-negative sepsis |  |
| A41.8 | Other specified sepsis | 1302246 |
| A41.81 | Sepsis due to Enterococcus |  |
| A41.9 | Sepsis, unspecified | 1302247 |
| R57.2 | Septic shock | 1313568 |
| R65.0 | Systemic Inflammatory Response Syndrome of infectious origin without organ failure | 1313599 |
| R65.1 | Systemic Inflammatory Response Syndrome of infectious origin with organ failure | 1313600 |
| R65.20 | Severe sepsis without septic shock |  |
| R65.21 | Severe sepsis with septic shock |  |
| R78.81 | Bacteraemia |  |

**Table S4. Conditions and weighting used to construct the Cambridge multimorbidity score**

|  | **Condition** | **Condition flags WSIC primary care database** | **Weight for mortality** |
| --- | --- | --- | --- |
| 1 | Hypertension | Hypertension | -2.09 |
| 2 | Anxiety/depression | Anxiety, Depression | 7.04 |
| 3 | Painful condition | Myalgic encephalomyelitis, Multiple sclerosis | 16.46 |
| 4 | Hearing loss | n/a | -3.94 |
| 5 | Irritable bowel syndrome | n/a | -1.33 |
| 6 | Asthma | Asthma | -2.73 |
| 7 | Diabetes mellitus | Diabetes | 10.23 |
| 8 | Coronary heart disease | Coronary heart disease (CHD) | 4.22 |
| 9 | Chronic kidney disease | Chronic kidney disease (CKD) | 16.61 |
| 10 | Atrial fibrillation | Atrial fibrillation | 22.14 |
| 11 | Constipation | n/a | 35.42 |
| 12 | Stroke and TIA | Stroke / transient ischemic attack | 20.63 |
| 13 | COPD | Chronic obstructive pulmonary disease (COPD) | 42.50 |
| 14 | Connective tissue disorder | Rheumatoid arthritis | -0.39 |
| 15 | Cancer | Cancer | 62.00 |
| 16 | Alcohol problems | n/a | 12.72 |
| 17 | Heart failure | Heart failure | 43.47 |
| 18 | Dementia | Dementia | 124.42 |
| 19 | Psychosis/bipolar disorder | Mental health | 7.20 |
| 20 | Epilepsy | Epilepsy | 18.26 |

**Table S5. Changes in urinary tract infection (UTI) incidence per 100,000 population per month during different stages of the COVID-19 pandemic**

|  | **Change in UTI incidence per 100,000 population per month** | **P-value** | **95% CI** | |
| --- | --- | --- | --- | --- |
|  |  |  | **Lower** | **Upper** |
| Pre-pandemic | -0.223 | 0.578 | -1.025 | 0.579 |
| Wave 1 | -24.227 | 0.000 | -28.431 | -20.023 |
| Between wave 1 and 2 | 17.570 | 0.000 | 14.340 | 20.801 |
| Wave 2 | -2.466 | 0.765 | -18.898 | 13.966 |
| Between wave 2 and 3 | 3.612 | 0.855 | -35.629 | 42.854 |
| Wave 3 | -7.409 | 0.000 | -8.208 | -6.610 |

**Table S6.** **Distribution of urinary tract infection (UTI) episodes related to antibiotic treatment**

|  | **Treated with antibiotics** | | **Not treated with antibiotics** | | **Total** | | **χ2 /** **H test** |
| --- | --- | --- | --- | --- | --- | --- | --- |
|  | **N = 190,993** | | **N = 184,866** | | **N = 375,859** | |  |
|  | **n** | **%** | **n** | **%** |  |  |  |
| **Mean age (years) (SD)** | 42.5 (23.0) | | 47.5 (23.0) | | 45.0 (23.0) | |  |
| **Age group** | | | | | | | |
| Under 16 | 13,262 | 6.9% | 10,573 | 5.7% | 23,835 | 6.3% | 0.000 |
| 16 - 64 | 141,370 | 74.0% | 122,260 | 66.1% | 263,630 | 70.1% |  |
| Above 64 | 36,361 | 19.0% | 52,033 | 28.1% | 88,394 | 23.5% |  |
| **Ethnicity** | | | | | | | |
| Non-white | 85,017 | 44.4% | 78,482 | 42.5% | 163,365 | 43.5% | 0.000 |
| White | 94,681 | 49.6% | 98,479 | 53.3% | 193,239 | 51.4% |  |
| Unknown | 11,244 | 5.9% | 7,905 | 4.3% | 19,255 | 5.1% |  |
| **Deprivation** | | | | | | | |
| 1 (most) | 20,022 | 10.5% | 19,513 | 10.6% | 39,535 | 10.5% | 0.000 |
| 2 | 46,633 | 24.4% | 44,590 | 24.1% | 91,223 | 24.3% |  |
| 3 | 36,873 | 19.3% | 35,812 | 19.4% | 72,685 | 19.3% |  |
| 4 | 20,981 | 11.0% | 20,378 | 11.0% | 41,359 | 11.0% |  |
| 5 (least) | 10,527 | 5.5% | 11,915 | 6.4% | 22,442 | 6.0% |  |
| Unknown | 55,957 | 29.3% | 52,658 | 28.5% | 108,615 | 28.9% |  |
| **Residential status** | | | | | | | |
| Occurred in care home | 1,957 | 1.0% | 3,374 | 1.8% | 5,331 | 1.4% | 0.000 |
| Not occurred in care home | 189,036 | 99.0% | 181,492 | 98.2% | 370,528 | 98.6% |  |
| **Co-morbidity** | | | | | | | |
| Mean Cambridge Multimorbidity Score (SD) | 0.1 (3.2) | | 0.3 (3.2) | | 0.2 (3.2) | |  |
| Number of long-term conditions: 0 | 99,575 | 52.1% | 77,181 | 41.7% | 176,756 | 47.0% | 0.000 |
| Number of long-term conditions: 1 | 40,045 | 21.0% | 39,873 | 21.6% | 79,918 | 21.3% |  |
| Number of long-term conditions: 2 and above | 51,373 | 26.9% | 67,812 | 36.7% | 119,185 | 31.7% |  |
| **Recent hospital admission** | | | | | | | |
| Hospital admission within 30 days prior to UTI | 1,163 | 0.6% | 1,621 | 0.9% | 2,784 | 0.7% | 0.000 |
| No admission within 30 days prior to UTI | 189,830 | 99.4% | 168,649 | 99.1% | 373,075 | 99.3% |  |
| **Pregnancy** | | | | | | | |
| Pregnant when UTI was diagnosed | 787 | 0.4% | 534 | 0.3% | 1,321 | 0.4% | 0.000 |
| Not pregnant when UTI was diagnosed | 190,206 | 99.6% | 184,332 | 99.7% | 374,538 | 99.6% |  |
| **Laboratory confirmed SARS-CoV-2 infection** | | | | | | | |
| Had positive results | 10,433 | 5.5% | 12,646 | 6.8% | 23,079 | 6.1% | 0.000 |
| No positive result | 180,560 | 94.5% | 172,220 | 93.2% | 352,780 | 93.9% |  |
| **Consultation method** | | | | | | | |
| Face-to-face | 124,449 | 65.2% | 99,624 | 53.9% | 224,073 | 59.6% | 0.351 |
| Remote | 48,451 | 25.4% | 41,592 | 22.5% | 90,043 | 24.0% |  |
| Unknown | 18,093 | 9.5% | 43,650 | 23.6% | 61,743 | 16.4% |  |
| **Documentation of diagnostic test as per NICE guidelines** | | | | | | | |
| In line | 41,369 | 21.7% | 16,217 | 8.8% | 57,586 | 15.3% | 0.000 |
| Not in line | 149,624 | 78.3% | 168,649 | 91.2% | 318,273 | 84.7% |  |
| **Outcome** | | | | | | | |
| 60-day bacteraemia, bloodstream infection, or sepsis | 418 | 0.2% | 1,539 | 0.8% | 1,957 | 0.5% | 0.000 |
| 60-day all-cause mortality | 737 | 0.4% | 850 | 0.5% | 1,587 | 0.4% |  |

**Table S7. Distribution of antibiotic prescriptions for a urinary tract infection (UTI)**

|  |  | **Recommended duration (days)** | **Number of prescriptions (n)** | **Number of non-prophylactic acute prescriptions (n, %)** | **Prescriptions with excess duration** | | | |
| --- | --- | --- | --- | --- | --- | --- | --- | --- |
|  |  |  |  |  | **(n)** | **(%)** | **95% CI, lower** | **95% CI, upper** |
| Nitrofurantoin  (n = 146,581) | ≥16 years, not pregnant | 3 | 144,174 | 105,149 (72.9%) | 52,991 | 50.4% | 50.1% | 50.7% |
|  | ≥ 12 years, pregnant | 7 | 400 | - | - | - | - | - |
|  | < 16 years | 3 | 2,007 | 1,561 (77.8%) | 697 | 44.7% | 42.2% | 47.2% |
| Trimethoprim  (n = 48,975) | ≥16 years, not pregnant | 3 | 39,208 | 29,695 (75.7%) | 15,999 | 53.9% | 53.3 | 54.5 |
|  | ≥ 12 years, pregnant | - | 17 | - | - | - | - | - |
|  | < 16 years | 3 | 9,750 | 8,389 (86.0%) | 4,575 | 54.5% | 53.5% | 55.6% |
| Amoxicillin  (n = 14,574) | ≥16 years, not pregnant | - | 11,743 | - | - | - | - | - |
|  | ≥ 12 years, pregnant | 7 | 275 | - | - | - | - | - |
|  | < 16 years | 3 | 2,556 | 2,268 (88.7%) | 2,116 | 93.3% | 92.2% | 94.3% |
| Cefalexin  (n = 13,375) | ≥16 years, not pregnant | - | 11,857 | - | - | - | - | - |
|  | ≥ 12 years, pregnant | 7 | 236 | - | - | - | - | - |
|  | < 16 years | 3 | 1,282 | 1,032 (80.5%) | 962 | 93.2 | 91.5 | 94.7 |

**Table S8. Changes in urinary tract infection (UTI) incidence per 100,000 population per month during different stages of the COVID-19 pandemic**

|  | **Odds ratio of experiencing bacteraemia, bloodstream infection, or sepsis** | **P-value** | **95% CI** | |
| --- | --- | --- | --- | --- |
|  |  |  | **Lower** | **Upper** |
| Pre-pandemic | Ref |  |  |  |
| Wave 1 | 1.02275 | 0.855 | 0.80403 | 1.30095 |
| Between wave 1 and 2 | 0.82563 | 0.056 | 0.67807 | 1.00531 |
| Wave 2 | 0.91008 | 0.364 | 0.74269 | 1.11520 |
| Between wave 2 and 3 | 1.16619 | 0.094 | 0.97396 | 1.39636 |
| Wave 3 | 0.77034 | 0.005 | 0.64138 | 0.92524 |

**Figure S1. Percentage of urinary tract infection (UTI) episodes with bacteraemia, bloodstream infection, or sepsis diagnosed within the following 60 days**

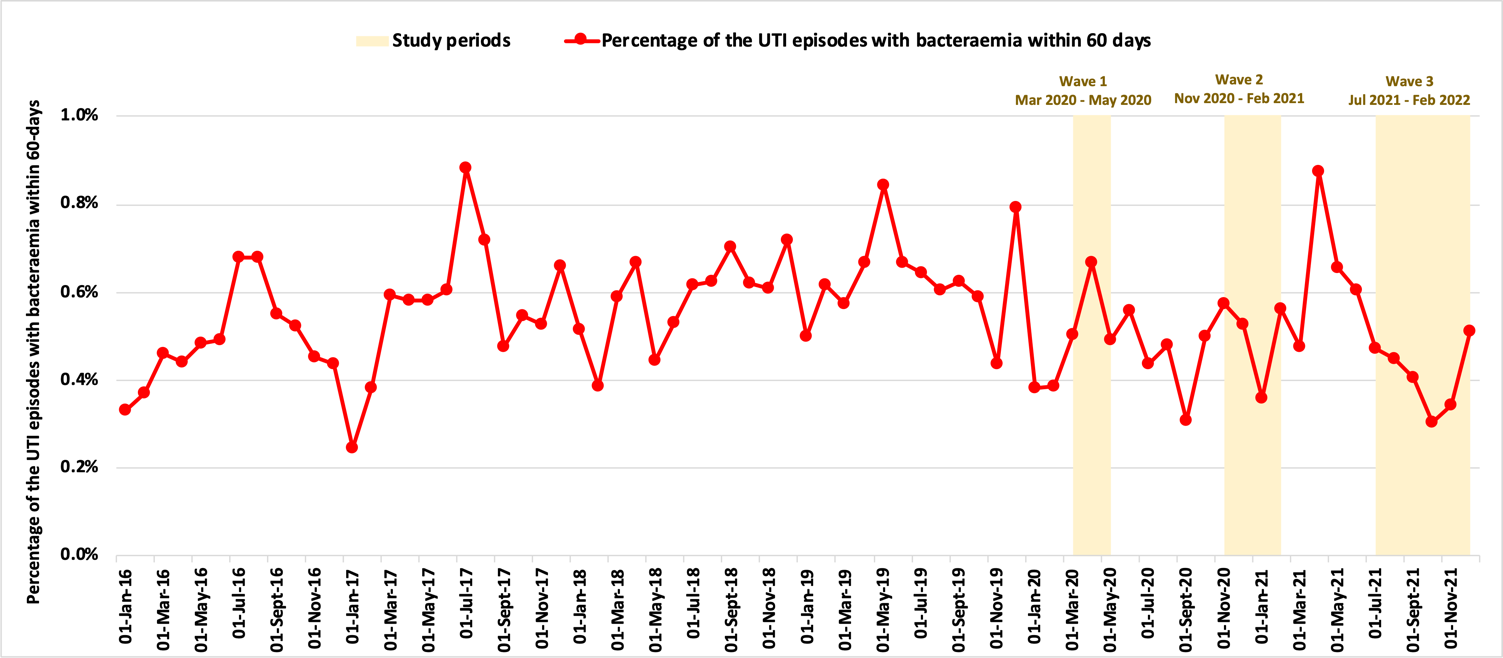
